# SIR-simulation of Corona pandemic dynamics in Europe

**DOI:** 10.1101/2020.04.22.20075135

**Authors:** Igor Nesteruk

## Abstract

The SIR (susceptible-infected-removed) model, statistical approach to the parameter identification and the official WHO daily data about the confirmed cumulative number of cases were used to estimate the characteristics of COVID-19 pandemic in Italy, Spain, Germany, France, Austria and Moldova. The final sizes and durations of epidemic outbreaks in these countries are calculated.

## Introduction

Here we consider the development of epidemic outbreak in Italy, Spain, Germany, France, Austria and the Republic of Moldova caused by coronavirus COVID-19 (2019-nCoV) (see e.g., [1]). For an epidemic of an infectious disease, the SIR model, connecting the number of susceptible *S*, infected and spreading the infection *I* and removed *R* persons, can be used [2-4]. The unknown parameters of this model can be estimated with the use of the cumulative number of cases *V=I+R* and the statistics-based method of parameter identification developed in [5, 6].

This approach was used in [6-12] to estimate the Corona pandemic dynamics in China, Republic of Korea, Italy, Austria and Ukraine. Usually the number of cases registered during the initial period of an epidemic is not reliable, since many infected persons are not detected. That is why the correct estimations of epidemic parameters can be done with the use of data sets obtained for later periods of the epidemic when the number of detected cases is closer to the real one. This fact necessitates a periodic reassessment of the epidemic’s characteristics and forecasts for its final size and duration. In this paper we will recalculate the pandemic parameters for Italy and Austria, provide estimations for Spain, Germany, France and Moldova, and compare with the further pandemic development.

### Data

The official data about the accumulated numbers of confirmed COVID-19 cases *V*_*j*_ in Italy, Spain, Germany, France, Austria and in the republic of Moldova from WHO daily situation reports (numbers 33-87), [1] are presented in Tables 1 and 2. The corresponding moments of time *t*_*j*_ (measured in days) are also shown in these tables. The data sets presented in Table 1 were used only for comparison with corresponding SIR curves. Table 2 was used for calculations and verifications of predictions.

**Table 1.** Official cumulative numbers of confirmed cases in Italy, Spain, Germany, France, Austria and Moldova during the initial stage of epidemics used only for comparison with SIR curves, [1]

| Day in<br>February<br>and<br>March,<br>2020 | Time<br>moments<br>In days $t_j$ | Number of<br>cases in Italy | Number of<br>cases in Spain | Number of<br>cases in<br>Germany | Number of<br>cases in<br>France | Number of<br>cases in<br>Austria | Number of<br>cases in the<br>Republic of<br>Moldova |
| --- | --- | --- | --- | --- | --- | --- | --- |
| 21 | -1 | 9 | 2 | 16 | 12 | 0 | 0 |
| 22 | 0 | 76 | 2 | 16 | 12 | 0 | 0 |
| 23 | 1 | 124 | 2 | 16 | 12 | 0 | 0 |
| 24 | 2 | 229 | 2 | 16 | 12 | 0 | 0 |
| 25 | 3 | 322 | 2 | 18 | 12 | 2 | 0 |
| 26 | 4 | 400 | 12 | 21 | 18 | 2 | 0 |
| 27 | 5 | 650 | 25 | 26 | 38 | 4 | 0 |
| 28 | 6 | 888 | 32 | 57 | 57 | 5 | 0 |
| 29 | 7 | 1128 | 45 | 57 | 100 | 10 | 0 |
| 1 | 8 | 1689 | 45 | 129 | 100 | 10 | 0 |
| 2 | 9 | 2036 | 114 | 157 | 191 | 18 | 0 |
| 3 | 10 | 2502 | 151 | 196 | 212 | 24 | 0 |
| 4 | 11 | 3089 | 198 | 262 | 282 | 37 | 0 |
| 5 | 12 | 3858 | 257 | 534 | 420 | 47 | 0 |
| 6 | 13 | 4636 | 374 | 639 | 613 | 66 | 0 |
| 7 | 14 | 5883 | 430 | 795 | 706 | 104 | 1 |
| 8 | 15 | 7375 | 589 | 1112 | 1116 | 112 | 1 |
| 9 | 16 | 9172 | 1024 | 1139 | 1402 | 131 | 1 |
| 10 | 17 | 10149 | 1639 | 1296 | 1774 | 182 | 3 |
| 11 | 18 | 12462 | 2140 | 1567 | 2269 | 302 | 4 |
| 12 | 19 | 15113 | 2965 | 2369 | 2860 | 361 | 4 |
| 13 | 20 | 17660 | 4231 | 3062 | 3640 | 504 | 8 |
| 14 | 21 | 21157 | 5753 | 3795 | 4469 | 800 | 12 |
| 15 | 22 | 24747 | 7753 | 4838 | 5380 | 959 | 23 |
| 16 | 23 | 27980 | 9191 | 6012 | 6573 | 1132 | 29 |
| 17 | 24 | 31506 | 11178 | 7156 | 7652 | 1332 | 30 |
| 18 | 25 | 35713 | 13716 | 8198 | 9043 | 1646 | 36 |
| 19 | 26 | 41035 | 17147 | 10999 | 10877 | 1843 | 49 |
| 20 | 27 | 47021 | 19980 | 18323 | 12475 | 2649 | 66 |
| 21 | 28 | 53578 | 24926 | 21463 | 14296 | 3024 | 80 |
| 22 | 29 | 59138 | 28572 | 24774 | 15821 | 3631 | 94 |
| 23 | 30 | 63927 | 33089 | 29212 | 19615 | 4486 | 109 |
| 24 | 31 | 69176 | 39673 | 31554 | 22025 | 5282 | 125 |
| 25 | 32 | 74386 | 47610 | 36508 | 24920 | 5888 | 149 |
| 26 | 33 | 80539 | 56188 | 42288 | 28786 | 7029 | 177 |
| 27 | 34 | 86498 | 64059 | 48582 | 32542 | 7697 | 199 |

**Table 2.** Official cumulative numbers of confirmed cases in Italy, Spain, Germany, France, Austria and Moldova used for calculations and verifications of predictions, [1]

| Day in<br>March<br>and<br>April,<br>2020 | Time<br>moments<br>in days $t_j$ | Number of<br>cases in Italy | Number of<br>cases in Spain | Number of<br>cases in<br>Germany | Number of<br>cases in<br>France | Number of<br>cases in<br>Austria | Number of<br>cases in the<br>Republic of<br>Moldova |
| --- | --- | --- | --- | --- | --- | --- | --- |
| 28 | 35 | 92472 | 72248 | 52547 | 37145 | 8291 | 231 |
| 29 | 36 | 97689 | 78797 | 57298 | 39642 | 8813 | 263 |
| 30 | 37 | 101739 | 85195 | 61913 | 43977 | 9618 | 298 |
| 31 | 38 | 105792 | 94417 | 67366 | 51477 | 10182 | 353 |
| 1 | 39 | 110574 | 102136 | 73522 | 56261 | 10711 | 423 |
| 2 | 40 | 115242 | 110238 | 79696 | 58327 | 11129 | 591 |
| 3 | 41 | 119827 | 117710 | 85778 | 63536 | 11525 | 591 |
| 4 | 42 | 124632 | 124736 | 91714 | 67757 | 11766 | 752 |
| 5 | 43 | 128948 | 130759 | 95391 | 69607 | 11983 | 864 |
| 6 | 44 | 132547 | 135032 | 99225 | 73488 | 12297 | 965 |
| 7 | 45 | 135586 | 140510 | 103228 | 77226 | 12640 | 1056 |
| 8 | 46 | 139422 | 146690 | 108202 | 81095 | 12969 | 1174 |
| 9 | 47 | 143626 | 152446 | 113525 | 85351 | 13248 | 1289 |
| 10 | 48 | 147577 | 157022 | 117658 | 89683 | 13560 | 1438 |
| 11 | 49 | 152271 | 161852 | 120479 | 92787 | 13807 | 1560 |
| 12 | 50 | 156363 | 166019 | 123016 | 94382 | 13937 | 1662 |
| 13 | 51 | 159516 | 169496 | 125098 | 97050 | 14043 | 1712 |
| 14 | 52 | 162488 | 172541 | 127584 | 102533 | 14234 | 1934 |
| 15 | 53 | 165155 | 177633 | 130450 | 105155 | 14370 | 2049 |
| 16 | 54 | 168941 | 182816 | 133830 | 107778 | 14448 | 2154 |
| 17 | 55 | 172434 | 188068 | 137439 | 108163 | 14603 | 2264 |
| 18 | 56 | 175925 | 191726 | 139897 | 110721 | 14662 | 2351 |
| 19 | 57 | 178972 | 195944 | 141672 | 111463 | 14710 | 2472 |
| 20 | 58 | 181228 | 200210 | 143457 | 113513 | 14783 | 2548 |

### SIR model

The SIR model for an infectious disease [2-5] relates the number of susceptible persons *S* (persons who are sensitive to the pathogen and **not protected**); the number of infected is *I* (persons who are sick and **spread the infection**; please don’t confuse with the number of still ill persons, so known active cases) and the number of removed *R* (persons who **no longer spread the infection**; this number is the sum of isolated, recovered, dead, and infected people who left the region); *α* and *ρ* are constants.

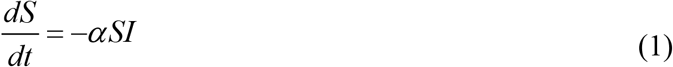

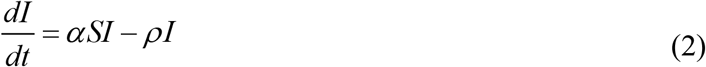

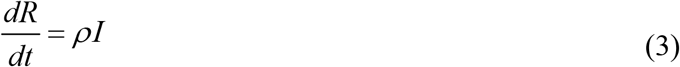

To determine the initial conditions for the set of equations (1–3), let us suppose that at the moment of the epidemic outbreak *t*_0_, [5, 6]:

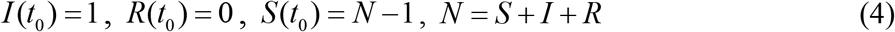

The analytical solution for the set of equations (1–3) was obtained by introducing the function *V* (*t*) = *I* (*t*) + *R*(*t*), corresponding to the number of victims or cumulative confirmed number of cases, [5, 6]:

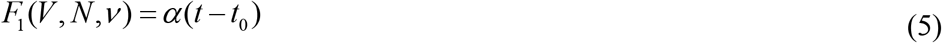

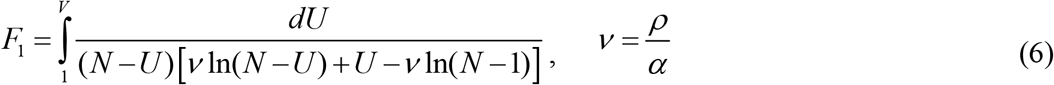

Thus, for every set of parameters *N, ν, α, t*_0_ and a fixed value of *V* the integral (6) can be calculated and the corresponding moment of time can be determined from (5). Then functions *I(t)* and *R(t)* can be easily calculated with the of formulas, [5, 6].

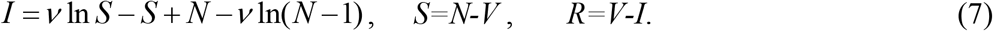

Function *I* has a maximum at *S* =*ν* and tends to zero at infinity, see [2, 3]. In comparison, the number of susceptible persons at infinity *S*_∞_ > 0, and can be calculated from the non-linear equation, [5, 6]:

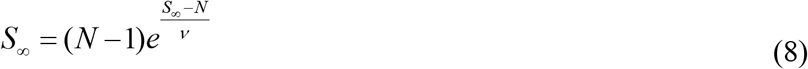

The final number of victims (final accumulated number of cases) can be calculated from:

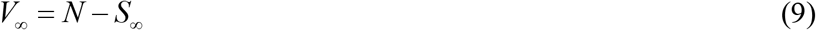

To estimate the duration of an epidemic outbreak, we can use the condition:

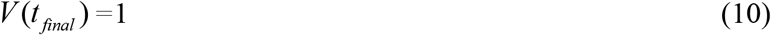

which means that at *t* > *t* _*final*_ less than one person still spread the infection.

### Parameter identification procedure

In the case of a new epidemic, the values of this independent four parameters are unknown and must be identified with the use of limited data sets. A statistical approach was developed in [5] and used in [6-12] to estimate the values of unknown parameters. The registered points for the number of victims *V*_*j*_ corresponding to the moments of time *t*_*j*_ can be used in order to calculate *F*_1 *j*_ = *F*_1_ (*V*_*j*_, *N,ν*) for every fixed values *N* and *ν* with the use of (6) and then to check how the registered points fit the straight line (5). For this purpose the linear regression can be used, e.g., [13], and the optimal straight line, minimizing the sum of squared distances between registered and theoretical points, can be defined. Thus we can find the optimal values of *α t*_0_ and calculate the, correlation coefficient *r*.

Then the F-test may be applied to check how the null hypothesis that says that the proposed linear relationship (5) fits the data set. The experimental value of the Fisher function can be calculated with the use of the formula:

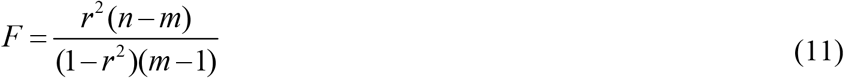

where *n* is the number of observations, *m*=2 is the number of parameters in the regression equation, [13]. The corresponding experimental value *F* has to be compared with the critical value *F*_*C*_ (*k*_1_, *k*_2_) of the Fisher function at a desired significance or confidence level (*k*_1_ = *m* −1, *k*_2_ = *n* − *m*), [14]. When the values *n* and *m* are fixed, the maximum of the Fisher function coincides with the maximum of the correlation coefficient. Therefore, to find the optimal values of parameters *N* and *ν*, we have to find the maximum of the correlation coefficient. To compare the reliability of different predictions (with different values of *n*) it is useful to use the ratio *F* / *F*_*C*_ (1, *n* − 2) at fixed significance level, [15]. We will use the level 0.001; corresponding values *F*_*C*_ (1, *n* − 2) can be taken from [14]. The most reliable prediction yields the highest *F* / *F*_*C*_ (1, *n* − 2) ratio.

## Results

Usually the number of cases during the initial period of an epidemic outbreak is not reliable. To avoid their influence on the results, only *V*_*j*_ values for the period March 28 – April 10, 2020 (35 ≤ *t* _*j*_ ≤ 48 ; *n*=14; *F*_*C*_ (1, *n* − 2) =18.6; see Table 2) were used to calculate the epidemic characteristics in every country. Since during the quarantine, the international people exchange is quite limited, we can apply the SIR model for every country assuming its parameters to be constant (but different for every country) during the fixed period of time. The results of calculations are shown in Tables 3 and 4. To illustrate the influence of data on the results of SIR simulations, other time periods: April 5 – 18, 2020 (43 ≤ *t* _*j*_ ≤ 56 ; *n*=14; *F*_*C*_ (1, *n* − 2) =18.6; France, Moldova) and March 29-April 18 (36 ≤ *t* _*j*_ ≤ 56 ; *n*=21; *F*_*C*_ (1, *n* − 2) =15.2; Italy, Spain) were used for second series of calculations for France and Moldova. The results are shown in Table 5.

**Table 3.** Epidemic characteristics for Italy, Spain and Germany. Optimal values of parameters, final sizes and durations (last two rows).

| Country | Italy, prediction 5 | Spain, prediction 1 | Germany |
| --- | --- | --- | --- |
| $N$ | 984556.8 | 736000 | 1023648 |
| $\nu$ | 882151.283536331 | 636763.648000000 | 946949.046049997 |
| $\alpha$ | 9.4016573087e-07 | 1.68916214450361e-06 | 1.9664814010e-06 |
| $t_0$ | -67.283729213432 | -22.8563373560822 | -22.835511732975 |
| $\rho$ | 0.8293684062279 | 1.07559704919762 | 1.86215768681056 |
| $1/\rho$ | 1.20573679017765 | 0.929716198780932 | 0.53701145025627 |
| $r$ | 0.9996573802724 | 0.999378854391215 | 0.99904312352174 |
| $F$, eq. (11) | 17503.1269829749 | 9650.57175588669 | 6261.40324472474 |
| $F / F_C(1, n-2)$ | 941.028332418004 | 518.847943864876 | 336.634583049717 |
| $S_\infty$, eq (8) | 787098 | 546870 | 874169.368888300 |
| $V_\infty$, eq (9) | 197459 | 189130 | 149479 |
| $t_{final}$, eq (10) | 148.5 | 105.6 | 104.8 |

**Table 4.** Epidemic characteristics for France, Austria and the Republic of Moldova. Optimal values of parameters, final sizes and durations (last two rows).

| Country | France, pred. 1 | Austria, pred. 4 | Moldova, pred. 1 |
| --- | --- | --- | --- |
| $N$ | 548182.4 | 75176.032 | 2911.36 |
| $\nu$ | 488472.457423422 | 67343.8674559792 | 1322.03320401920 |
| $\alpha$ | 2.601817507993e-06 | 1.924971386379e-05 | 0.0001386860412 |
| $t_0$ | -21.1949054853200 | -15.8059189609884 | 12.9079807669692 |
| $\rho$ | 1.27091619189685 | 1.29635017900866 | 0.18334755141242 |
| $1/\rho$ | 0.786833944185958 | 0.771396507049289 | 5.45412247012025 |
| $r$ | 0.996770038705090 | 0.996465578483694 | 0.99726948597323 |
| $F$, eq. (11) | 1848.61225180740 | 1688.59570093583 | 2188.39255686532 |
| $F / F_C(1, n-2)$ | 99.3877554735161 | 90.7847151040770 | 117.655513809963 |
| $S_\infty$, eq. (8) | 433257 | 60068 | 453 |
| $V_\infty$, eq. (9) | 114925 | 15108 | 2458 |
| $t_{final}$, eq. (10) | 106.2 | 86.5 | 109.9 |

**Table 5.** Results of second series of calculations with the use data from period April 5 – 18, 2020 for France and the Republic of Moldova, and March 29-April 18, 2020 for Italy and Spain. Optimal values of parameters, final sizes and durations (last two rows).

| Country | Spain, pred.2 | Italy, pred. 6 | France, pred. 2 | Moldova, pred. 2 |
| --- | --- | --- | --- | --- |
| $N$ | 760152,448 | 1125589,888 | 688720 | 6079,36 |
| $\nu$ | 643334,866338 | 1008518,7886 | 621554,6679951 | 4055,26811705 |
| $\alpha$ | 1,06816546e-06 | 6,759461602e-07 | 1,970170995e-06 | 7,641517427e-05 |
| $t_0$ | -43,8057685 | -88,26304932 | -29,7952392 | 4,3218081122 |
| $\rho$ | 0,6871880836 | 0,681704402658 | 1,2245689787 | 0,30988401 |
| $1/\rho$ | 1,455205676 | 1,46691145913 | 0,81661385959 | 3,2270137723 |
| $r$ | 0,997926053 | 0,99969602932 | 0,997453623886 | 0,998814863 |
| $F$, eq. (11) | 4566,3928836 | 31238,7652767 | 2347,293599 | 5053,707894 |
| $F/F_C(1, n-2)$ | 300,420584 | 2055,181926 | 126,1985806 | 271,7047254 |
| $S_\infty$, eq. (8) | 539142 | 899854 | 558897 | 2539 |
| $V_\infty$, eq. (9) | 221011 | 225736 | 129823 | 3541 |
| $t_{final}$, eq. (10) | 135.3 | 177.8 | 119.1 | 114.5 |

The SIR curves and markers representing the *V*_*j*_ values taken for calculations (“circles”); for comparisons (“triangles”) and verifications of predictions (“stars”) are shown in Figs. 1-6. For Italy, Spain, France and Moldova, the second sets of optimal parameters (from Table 5) were used to calculate SIR curves.

**Fig. 1.**
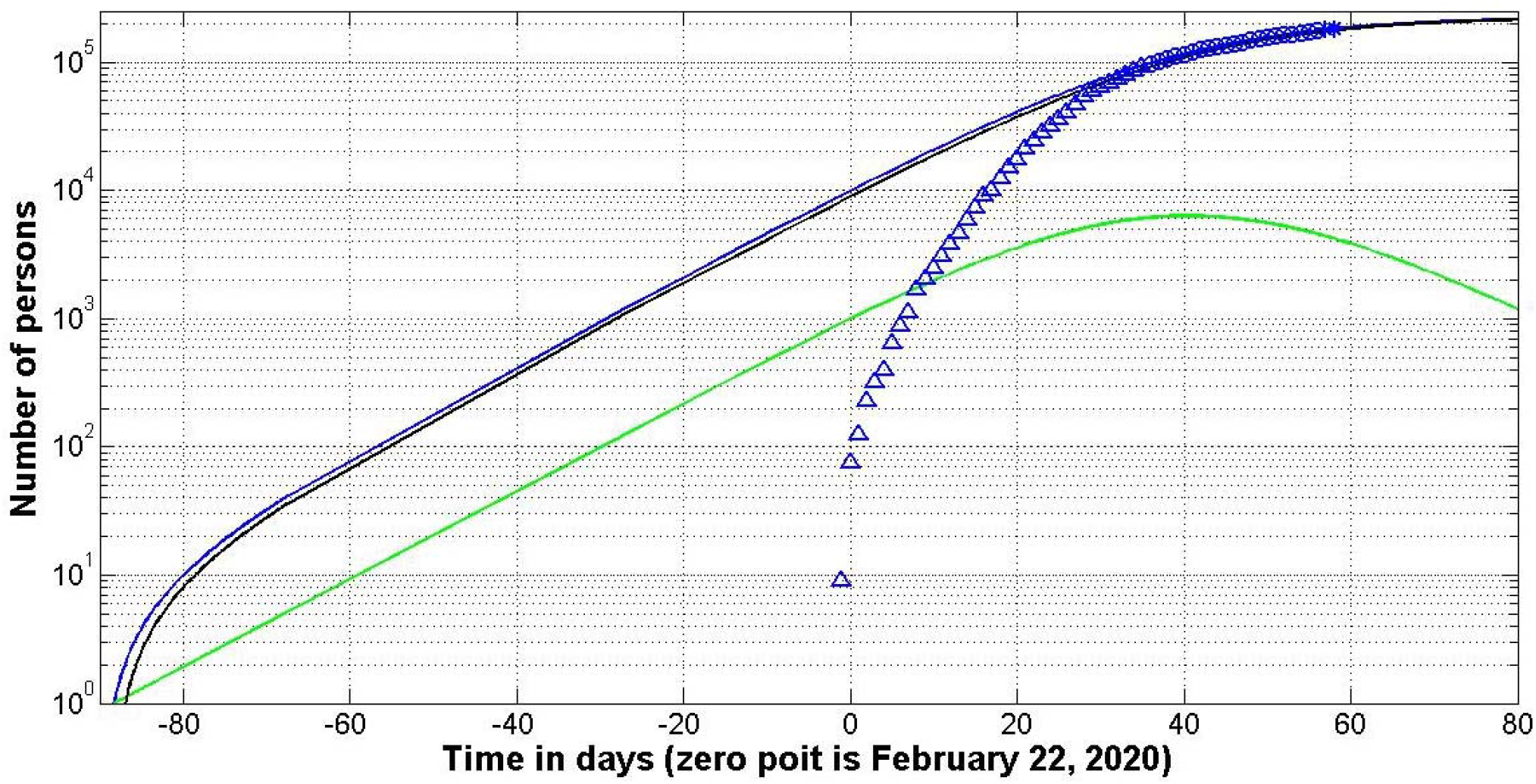
Italy, prediction 6: SIR curves (lines) and accumulated number of cases (markers) versus time. Numbers of infected *I* (green), removed *R* (black) and victims *V=I+R* (blue line).

It can be seen that the previous predictions for Italy and Austria [11] were more optimistic. Fresh data sets has showed that the final number of cases in Italy could reach 226,000 and their appearance can stop only after August 18, 2020 (see Table 5, prediction 6). The epidemic stop in Austria is expected after May 21, 2020 (see Table 4). These estimations are valid only when the quarantine measures, isolation rate and the coronavisus activity will be same as for the periods taken for calculations.

Tables 3-5 and Figs. 1, 2 illustrate that the estimations of parameter *t*_*0*_ values are very different from the results published in [11]. In particular, according to the prediction in Italy the first COVID-19 cases could happen even after November 27, 2019. This results correlates with the information form Giuseppe Remuzzi, director of the Mario Negri Institute for Pharmacological Research that “virus was circulating before we were aware of the outbreak in China”, [16]. Table 5 and Fig. 3 show that the epidemic outbreak in France could happen around January 25, 2020. This estimation correlates with the results of paper [17], where the first COVID-19 cases with 5 Chinese tourists are described. They were from Wuhan and arrived in Europe on January 17-22, 2020.

**Fig. 2.**
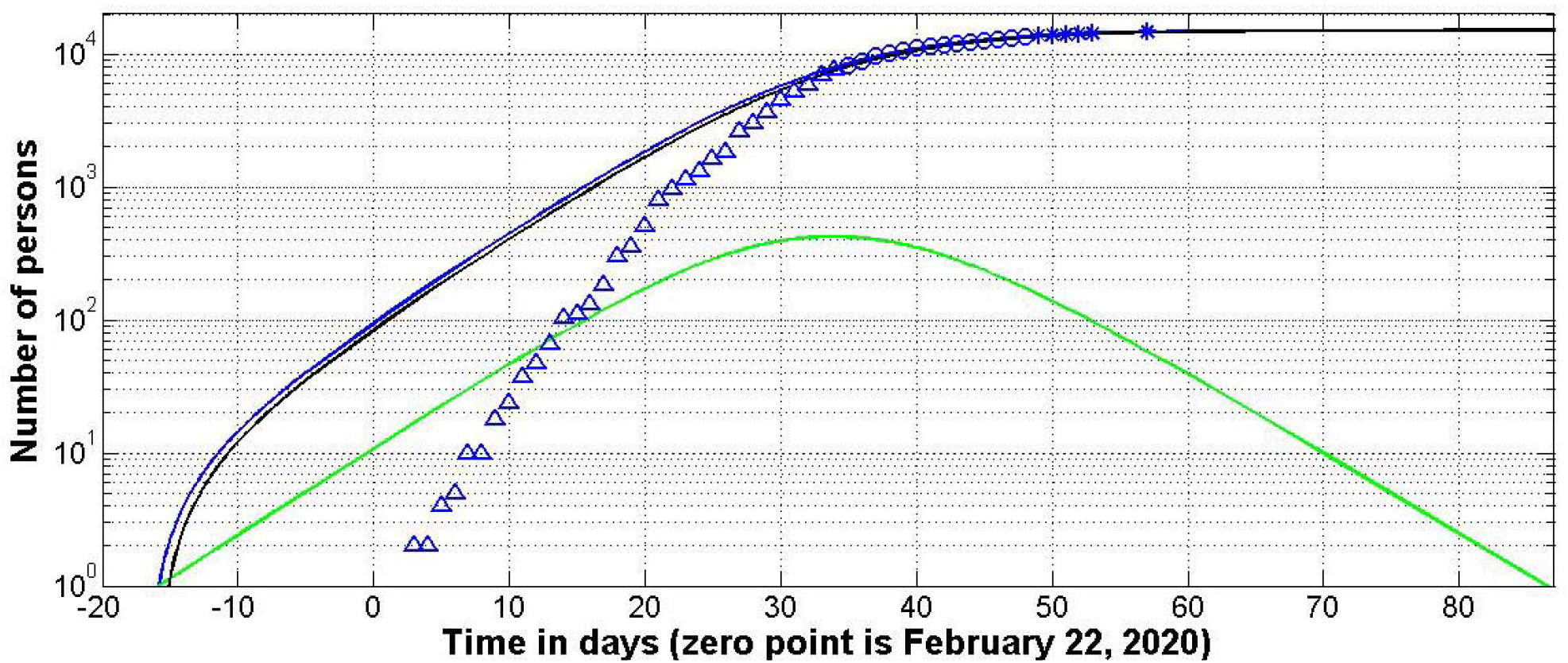
Austria, prediction 4: SIR curves (lines) and accumulated number of cases (markers) versus time. Numbers of infected *I* (green), removed *R* (black) and victims *V=I+R* (blue).

**Fig. 3.**
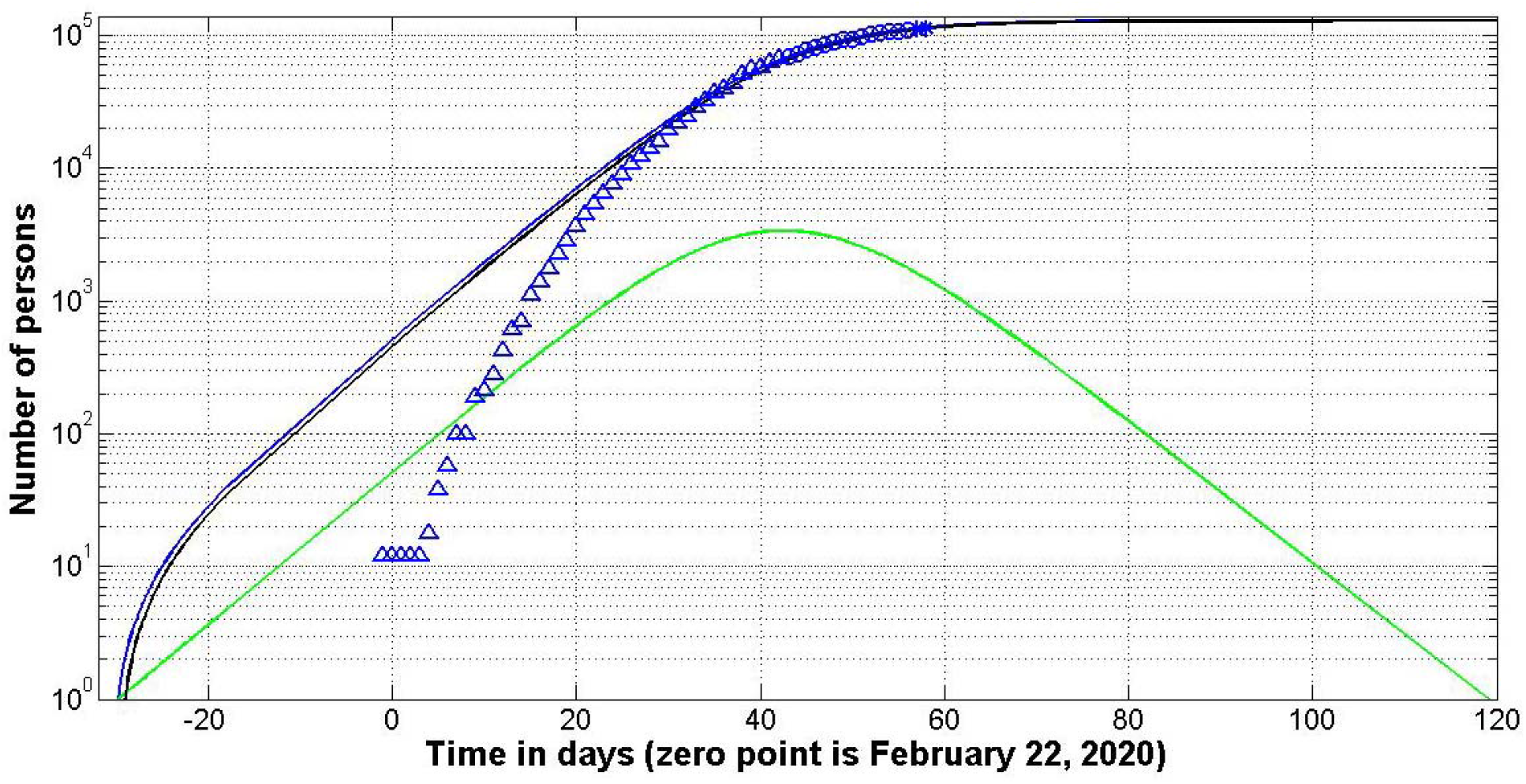
France, prediction 2: SIR curves (lines) and accumulated number of cases (markers) versus time. Numbers of infected *I* (green), removed *R* (black) and victims *V=I+R* (blue).

Fig. 4 illustrates that even second prediction for Spain looks too optimistic (“stars” deviate from the blue line more than for other countries). The second prediction for this country has lower value of *F* / *F*_*C*_ (1, *n* − 2) in comparison with the first one (see Tables 3 and 5), but for the first prediction the deviations are even larger. Probably the final size and the duration of the epidemic in Spain have to be re-estimated after obtaining new data.

**Fig. 4.**
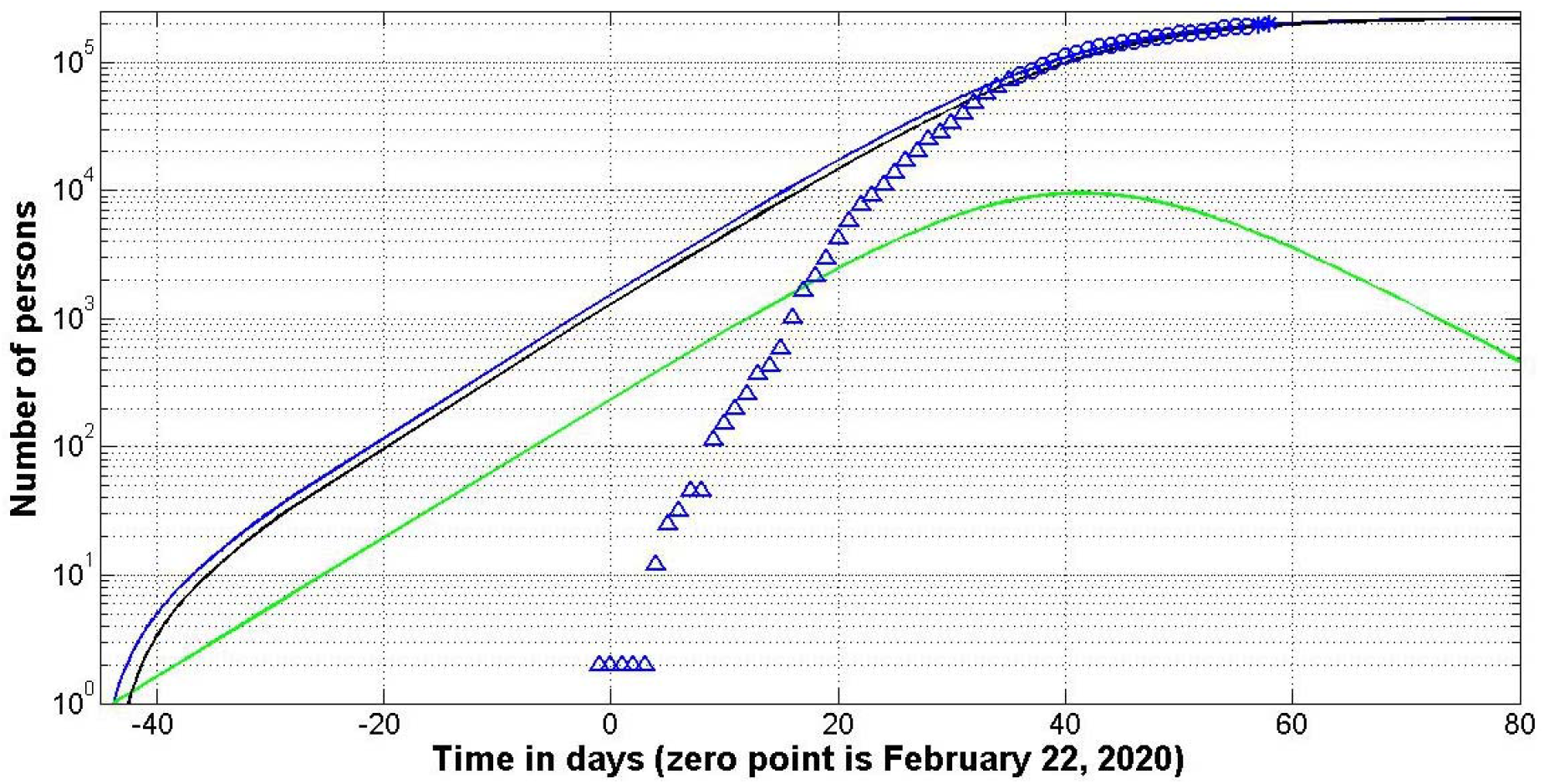
Spain, prediction 2: SIR curves (lines) and accumulated number of cases (markers) versus time. Numbers of infected *I* (green), removed *R* (black) and victims *V=I+R* (blue).

The *V* and *R* curves are very close in Fig. 5. It means that the time of spreading infection in Germany is very short (in comparison, for example, with Moldova, see Fig. 6). SIR model allows calculating the average time of spreading infection 1/ *ρ*. For Germany this value could be estimated as 0.54 days; for Austria and France approximately 0.8 days; for Italy and Spain – 1.5 days and 3.2 days for the Republic of Moldova (see Table 3-5). By comparison, in South Korea this time was approximately 4.3 hours, [8].

**Fig. 5.**
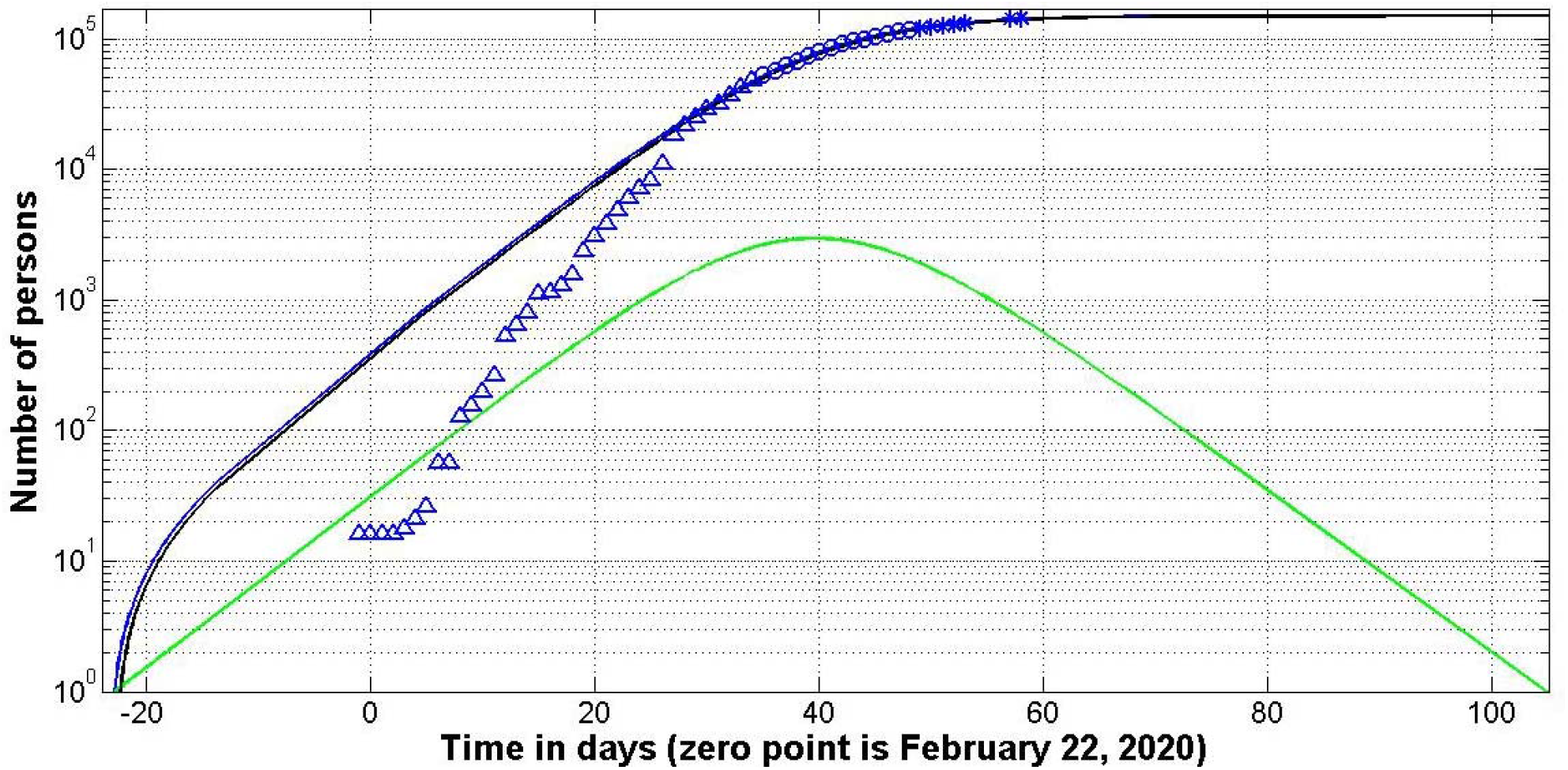
Germany: SIR curves (lines) and accumulated number of cases (markers) versus time. Numbers of infected *I* (green), removed *R* (black) and the number of victims *V=I+R* (blue line).

**Fig. 6.**
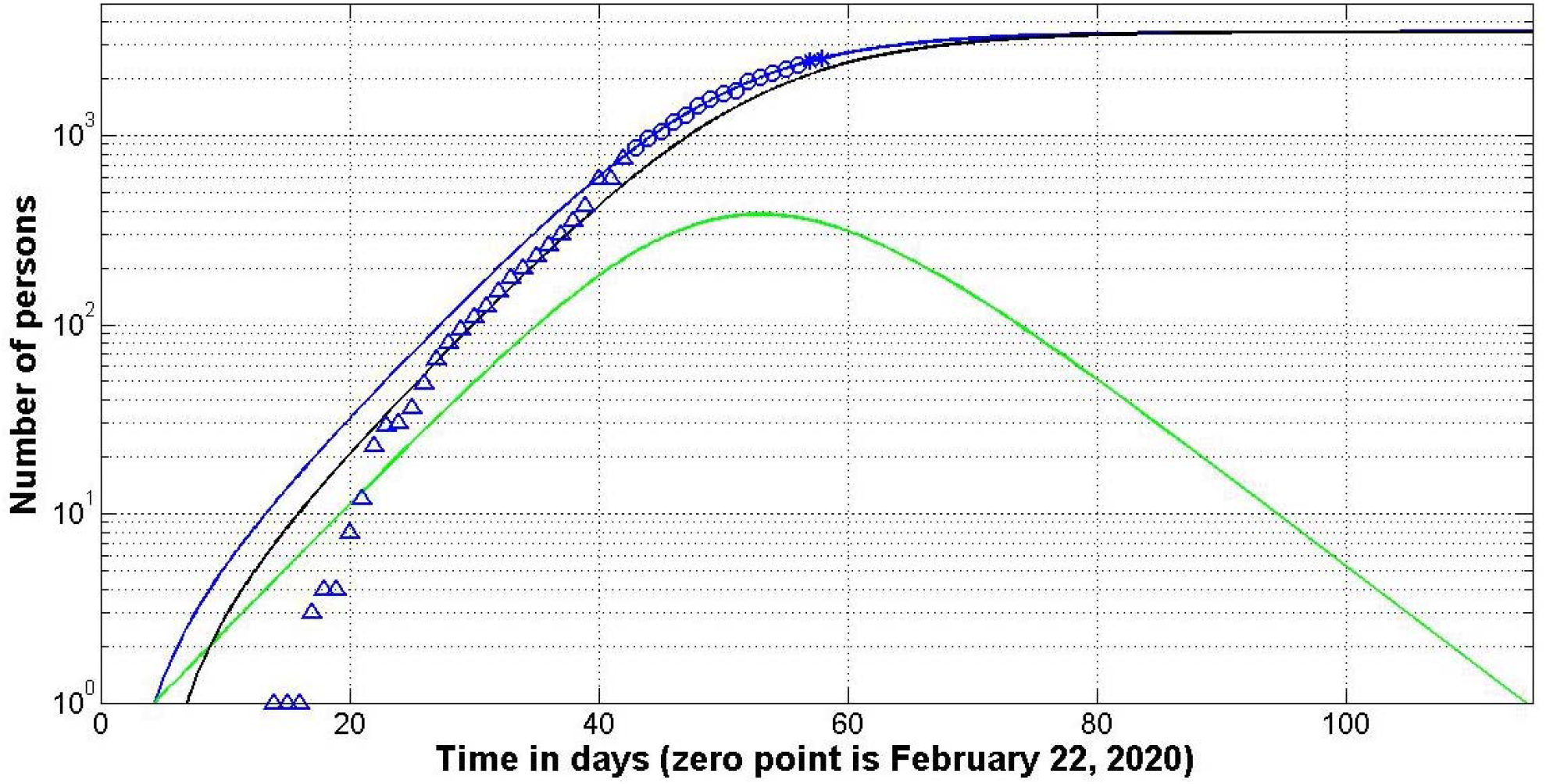
Moldova, prediction 2: SIR curves (lines) and accumulated number of cases (markers) versus time. Numbers of infected *I* (green), removed *R* (black) and victims *V=I+R* (blue line).

## Discussion

Figs. 1-6 illustrate that close to the epidemic outbreaks, the information about number of cases is not complete, since it is difficult to detect all the infected persons, especially those with mild illness. As a result, SIR simulation using data sets from the initial epidemic period has limited accuracy, and predictions of final size and duration are too optimistic (see [10-12]). The data quality may be very different and unpredictable. The only way to obtain the reliable results is to compare the *V(t)* curve with *V*_*j*_ data obtained after day of calculations. If the discrepancy after some days of observation is too large, new calculations must be performed with the use of fresh data. “Stars” in Figs. 1-3, 5,6 illustrate that the accuracy of calculations is good enough. Probably, the prediction for Spain is too optimistic and can be updated later.

It looks, that SIR model can determine the real time of epidemic outbreak. Due to the data incompleteness, the “hidden” period could be very long. For example, the real epidemic outbreaks in Italy and Spain probably happened in November, 2019 and January, 2020 respectively (see Table 5 and Figs. 1, 4). There is no reason to think that the “hidden” period in China was shorter. According to [18], the first laboratory confirmed case was recorded on December 8, 2019. Therefore the real “zero” patient could get infection in October-November, 2019 and probably had no connection with the Wuhan fish market. Chinese authorities notified WHO about the epidemic outbreak only on January 3, 2020 (see [18]). This delay and obstacles to the dissemination of truthful information (an example is doctor Li Wenliang) has led to tragic consequences in Europe and other parts of the world.

SIR curves could be useful to estimate the number of persons who are still spreading the infection, so known “hidden” patients (see green lines in Figs. 1-6). These information could be useful to plan the relaxation of quarantine measures. If medical and other services are able to quickly isolate persons who have contacted, for example, with 100 “hidden” patients, then there is no need to wait until the number of patients on the green curve reaches the value of unity. On the other hand, such attenuations may extend the time of occurrence of new cases and the number of deaths. A compromise between medicine and economic interests must be found here.

## Conclusions

The SIR (susceptible-infected-removed) model and statistical approach to the parameter are able to make some reliable estimations for the epidemic dynamics, e.g., the real time of the outbreak, final size and duration of the epidemic and the number of persons spreading the infection versus time. This information may be useful to regulate the quarantine activities and to predict the medical and economic consequences of the pandemic. Unfortunately, the number of patients in Europe already exceeds one million. The risk of catching the infection will persist until at least mid-August, 2020. Such fatal consequences could have been avoided if timely and truthful information came from China.

## Data Availability

Data is in text

## Acknowledgements

I would like to express my sincere thanks to Gerhard Demelmair and Ihor Kudybyn for their help in collecting and processing data.

